# Routine Intracoronary Imaging-guided Left Main Coronary Intervention

**DOI:** 10.1101/2024.02.09.24302611

**Authors:** Yoshinobu Murasato, Hitoshi Nakashima, Hiroshi Sugino, Masaya Arikawa, Fumiaki Mori, Yasunori Ueda, Keisuke Matsumura, Mitsuru Abe, Tomomi Koizumi, Mitsuhiro Shimomura, Kazuteru Fujimoto, Takahiro Saeki, Shogo Imagawa, Takashi Takenaka, Yukiko Morita, Katsuro Kashima, Akira Takami, Yujiro Ono, Atsuki Fukae, Hisako Yoshida, LM-JANHO investigators

## Abstract

**Background:** Left main (LM) percutaneous coronary intervention (PCI) with routine intracoronary imaging guidance is recommended; however, its real-world effectiveness remains unclear. This study aimed to investigate the outcomes in a Japanese National Hospital Organization cohort in which routine imaging guidance was adopted.

**Methods:** Of the 806 consecutive patients undergoing drug-eluting stent implantation for de novo unprotected LM lesions across 19 hospitals, 743 were analyzed after excluding 63 owing to criteria mismatch or incomplete follow-up. The primary endpoint was 1-year major adverse cardiovascular and cerebrovascular events (MACCE), comprising all-cause death, cerebrovascular disorder, clinical-driven revascularization, and myocardial infarction.

**Results:** The cohort exhibited increased prevalences of diabetes mellitus, prior myocardial infarction, and prior revascularization. Acute coronary syndrome was present in 31.2% of the patients, with 39.3% classified as Canadian Cardiovascular Society functional angina (CCS) class ≥III. LM bifurcation lesions were observed in 78.0% of the patients, with two-stent implantation in 8.8% of the patients. MACCE occurred in 17.5% of the patients, with target lesion revascularization and cardiac death rates of 2.0% and 3.4%, respectively. Independent risk factors for MACCE included CCS class ≥III (hazard ratio [HR], 2.07), mechanical cardiac support device use (HR, 2.17), two-stent implantation (HR, 2.49), 10% increase in left ventricular ejection fraction (HR, 0.72), and radial access (HR, 0.62).

**Conclusion:** Routine imaging-guided LM-PCI is associated with a lower incidence of target lesion revascularization and cardiac death. However, severe left ventricular dysfunction and multiple-vessel involvement are associated with higher mortality and revascularization risks, requiring comprehensive management beyond imaging-guided PCI.

**Clinical Perspective:** *What is new?:*

- This study clarifies the clinical outcomes of left main coronary intervention guided by routine intracoronary imaging, revealing a low frequency of target lesion revascularization and cardiac death.
- Despite the favorable local efficacy of imaging guidance, patients with severe left ventricular dysfunction and multiple-vessel involvement still face elevated risks of mortality and revascularization.

*What are the clinical implications?:*

- Intracoronary imaging guidance in the left main coronary artery is crucial for optimizing intervention treatments and enhancing local efficacy at the treated sites.
- Despite these improvements, the high mortality rate associated with serious myocardial damage from left main coronary artery obstruction underscores the importance of careful consideration in such cases.
- Coronary artery disease involving the left main and multiple vessels carries an elevated risk of additional revascularizations beyond the target lesion, emphasizing the need for comprehensive management strategies.

## Introduction

Intracoronary imaging guidance is recommended for complex percutaneous coronary intervention (PCI) to reduce mortality and target lesion revascularization^1–8^. It is also recommended for left main (LM) PCI^9, 10^, which has a large perfusion territory and is associated with a higher risk of hemodynamic collapse or fatal events. Imaging guidance is associated with sufficient stent expansion with less malapposition owing to optimal device selection and swift detection of stent failure or deformation^11, 12^. Additionally, imaging-guided bifurcation intervention is useful for the prediction of side branch (SB) compromise^11^ and prevention of unnecessary aggressive SB treatments, such as two-stent deployment or oversized SB dilation, leading to overtreatment regardless of uncertain angiographic findings in the bifurcation^13^.

A meta-analysis of 11 randomized control trials comparing LM-PCI and coronary artery bypass graft (CABG), including 11,518 patients, demonstrated similar rates of all-cause mortality when the Synergy between Percutaneous Coronary Intervention with Taxus and Cardiac Surgery (SYNTAX) score was <32^14^. Therefore, the guidelines have been changed so that LM-PCI is ranked in class I treatment, similar to CABG, when the SYNTAX score is <22, which indicates that anatomical lesion complexity is low^9^.

Intravascular imaging guidance has become more popular in Japan than in other countries because of reimbursement of intracoronary imaging devices through social insurance. Intracoronary imaging guidance was used in 95% of all PCIs in the nationwide survey in 2014^15^. In the Japanese Circulation Society 2018 Guidelines on Revascularization of Stable Coronary Artery Disease, intravascular ultrasound (IVUS) guidance is strongly recommended as class I in LM-PCI, and optical coherence tomography (OCT)/optical frequency-dependent imaging (OFDI) guidance is also recommended as class IIa in bifurcation PCI^10^. However, most of the evidences are derived from small-scale randomized trials comparing imaging guidance and angio-guidance or propensity-match comparison in large-scale registries, in which more complex lesions are selected according to the cases treated under imaging guidance^3^. Therefore, the efficacy of routine intracoronary imaging for LM-PCI in daily practice has not yet been clarified. We aimed to investigate the clinical outcomes of LM-PCI in a large cohort of the Japanese National Hospital Organization (LM-JANHO) group, where intracoronary imaging guidance is routinely used.

## Methods

### Study population and design

The LM-JANHO is a multicenter, retrospective, observational registry study. In total, 806 consecutive patients who underwent LM-PCI with a drug-eluting stent (DES) between January 2016 and December 2020 were retrospectively enrolled from 19 institutes in the Japanese National Hospital Organization group.

The inclusion criteria were as follows: (1) de novo LM lesions including significant stenosis in the LM (≥50%) and/or daughter branches (≥75%) within a distance of 5 mm from the carina, which were treated with DES implantation in LM or crossover from LM to daughter branches; (2) suitable lesion for DES implantation in the LM; (3) patient age >20 years; and (4) tolerability of dual antiplatelet therapy for >6 months. The exclusion criteria were as follows: (1) in-stent restenosis lesions; (2) chronic total occlusion in the LM or adjusting branches; (3) the left anterior descending artery (LAD) and/or left circumflex artery (LCX) protected by prior CABG; (4) female with possible or definite pregnancy; (5) unsuitable candidate as judged by the responsible doctor, and (6) refusal to provide personal information for the study after receiving study information, in accordance with the opt-out system.

After the exclusion of 38 patients (inclusion criteria unmet in 26, lost to follow-up in 11, physician’s decision in one) and 25 patients with any deficit of 1-year follow-up data, the data of 743 patients were analyzed (Figure 1). The study protocol was approved by the Institutional Review Board of the Japanese National Hospital Organization and each attending hospital. This study was conducted using an opt-out system and was disclosed to the patients and requirement for informed consent was waived. The study protocol was developed in accordance with the Declaration of Helsinki and registered in the University Hospital Medical Information Network (ID: UMIN 000037332) prior to the initiation of enrollment.

**Figure 1.**
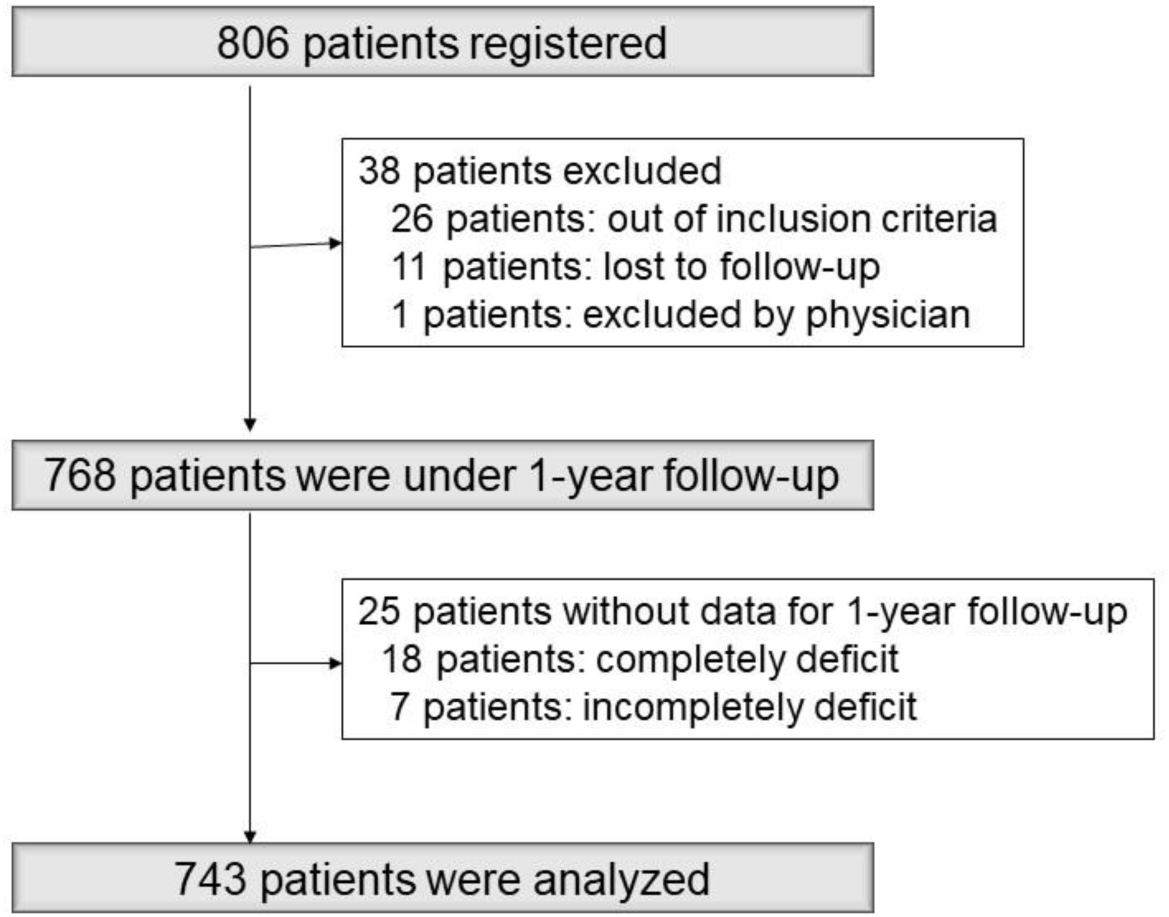
Study flow of the extraction of the analyzed patients.

### Percutaneous coronary intervention protocol

All patients were treated with sufficient periprocedural heparin and dual antiplatelet therapy with aspirin (100 mg) and either clopidogrel (50–75 mg) or prasugrel (3.75 mg), which was continued for at least 6 months. In cases with deficits in PCI initiation, the procedure was performed after administering the loading dose (aspirin 200 mg, clopidogrel 300 mg, and prasugrel 20 mg). Imaging was strongly recommended before PCI to assess the lesion and select the optimal therapeutic device and after the PCI procedure for its optimization. The use of IVUS or OCT/OFDI was at the operator’s discretion. If any stent failure (stent malapposition >400 μm, insufficient stent expansion [<80%] compared with the mean value of proximal and distal references, and stent deformation) or serious dissection in the SB or stent proximal and/or distal edges was observed on imaging, prompt additional treatment was performed. The proximal optimal technique (POT), using a short balloon according to the LM vessel size, was recommended; however, its use relied on the operator’s discretion after the imaging assessment for LM stent expansion. After LM crossover stenting to the LAD or LCX, opposite branch ostial dilation and the choice of method (kissing balloon inflation [KBI] or opposite branch dilation alone) were also dependent on the operator’s discretion.

### Data collection

Patient background, lesion characteristics, on-site visual assessment of coronary angiography, PCI procedure, medical treatment, and clinical outcome at 1-year follow-up were registered from each hospital to the REDCap system, a web-based application designed for streamlined data collection and management, in Osaka Metropolitan University Hospital center for clinical research and innovation.

In this study, acute coronary syndrome included acute myocardial infarction accompanied by elevated cardiac biomarker levels with or without ST elevation on electrocardiography and unstable angina, defined as new-onset or increased-severity angina within 1 month, resting angina, and postinfarction angina without elevated cardiac biomarker levels. Stable angina was defined as stable symptoms during high exertion, with no chest pain at rest in the past month. Silent myocardial ischemia was defined as the objective evidence of myocardial ischemia without evident chest symptoms. Cardiogenic shock was defined as a sustained episode of systolic blood pressure <80 mmHg and cardiac index <1.8 L/min⸱m^2^, determined to be secondary to cardiac dysfunction, and/or the requirement of a parenteral inotropic or vasopressor agent or mechanical support to maintain blood pressure and cardiac index within 24 h before PCI. Pulmonary edema was defined as symptoms of heart failure within 24 h before PCI, including dyspnea on mild activity, orthopnea, body fluid retention, moist rales, and neck vein distention, which were equivalent to Congestive Heart Failure in the New York Heart Association functional classification class IV. Cardiac death was defined as death due to cardiac diseases, including heart failure, fatal arrhythmia, and sudden death. Vascular death was defined as death due to a vascular disease, except for coronary artery disease, such as diseases of the cerebrovascular artery, aorta, and peripheral and pulmonary arteries. Target lesion revascularization (TLR) was defined as any revascularization by PCI or CABG in the treated LM and adjusting proximal LAD and LCX within 5 mm from the branch ostium. Target vessel revascularization (TVR) was defined as revascularization of the LM, LAD, or LCX. Clinically driven revascularization included TVR and non-TVR. Stent thrombosis included definite and probable stent thrombosis as defined by the Academic Research Consortium. Cerebrovascular disorders included stroke with any new-onset neurological deficit due to the occlusion of the cerebrovascular artery and cerebrovascular hemorrhage, except for traumatic hemorrhage. Successful PCI was defined as the achievement of Thrombolysis in Myocardial Infarction flow grade III with residual stenosis of ≤25% in the target lesion^16^.

### Endpoint

Major adverse cardiovascular and cerebrovascular events (MACCE) were defined as composite endpoints of the following at 1-year follow-up: all-cause death, cerebrovascular disorder, any clinically driven revascularization, and myocardial infarction.

### Statistical analyses

Continuous variables are expressed as mean±standard deviation. Categorical variables are presented as percentages. Cox proportional hazard models were constructed to evaluate the association between each patient background, lesion characteristics, PCI procedures, and MACCE. Hazard ratios (HRs) and 95% confidence intervals (CIs) were calculated. The rates of MACCE incidence between patient background, legion characteristics, PCI procedures were compared by estimating cumulative incidence functions against the days from PCI. All reported p-values were determined by two-sided analysis, and p-values <0.05 were considered significant. Analyses were performed using the R software (version 4.2.2) (R Foundation for Statistical Computing, Vienna, Austria).

## Results

### Baseline patient characteristics

Baseline patient characteristics are summarized in Table 1. Increased prevalence rates of male sex (78.7%), dyslipidemia (71.0%), diabetes mellitus (49.3%), prior myocardial infarction (25.2%), and prior PCI or CABG (43.6%) were observed. The mean left ventricular ejection fraction was 56±15%. Acute coronary syndrome was observed in 31.2% of the patients, and a severe hemodynamic collapse state (cardiogenic shock, cardiopulmonary arrest, and pulmonary edema) was observed in 2.4–6.1% of the patients. Canadian Cardiovascular Society (CCS) class ≥3 was observed in 39.3% of the patients.

**Table 1.**
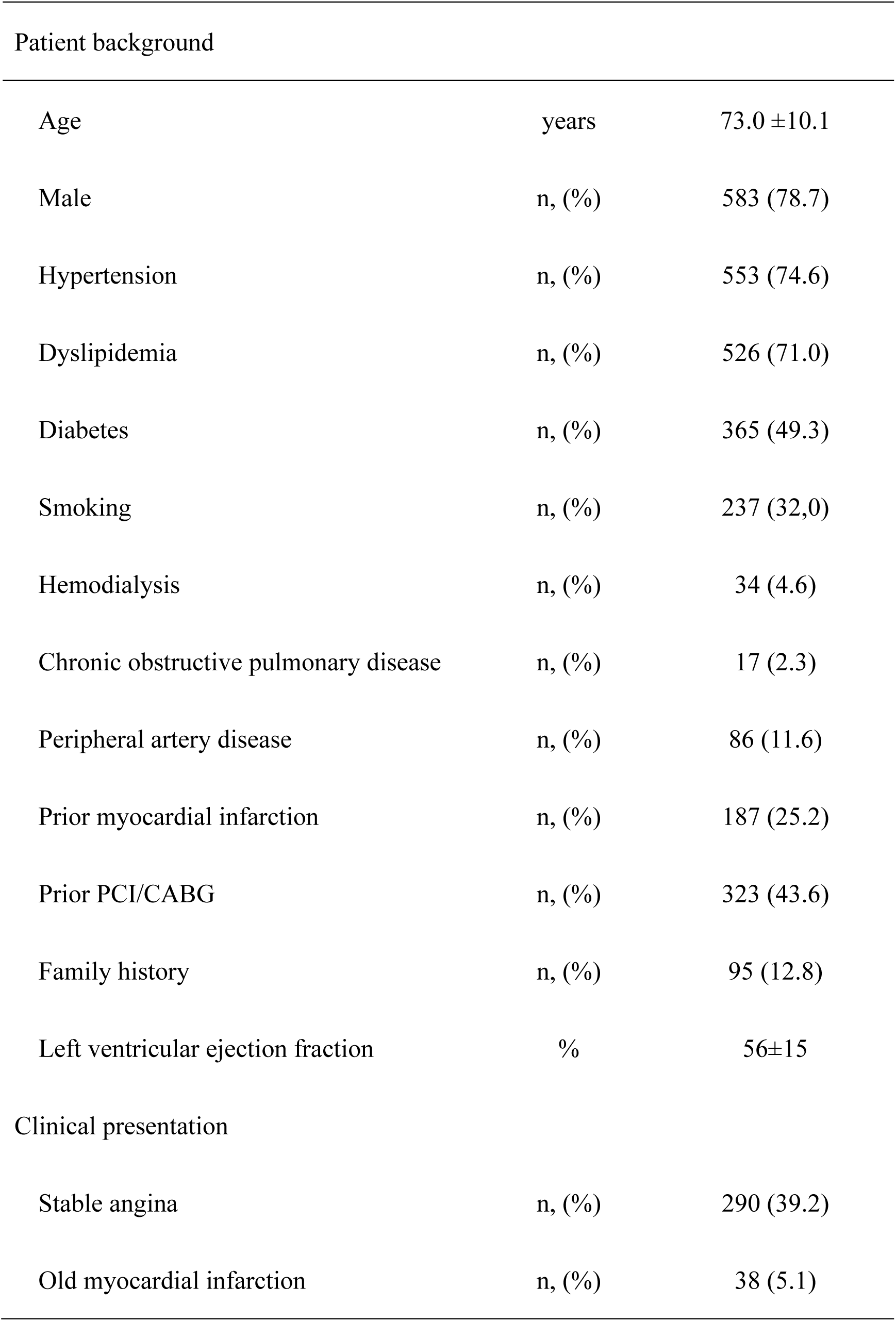
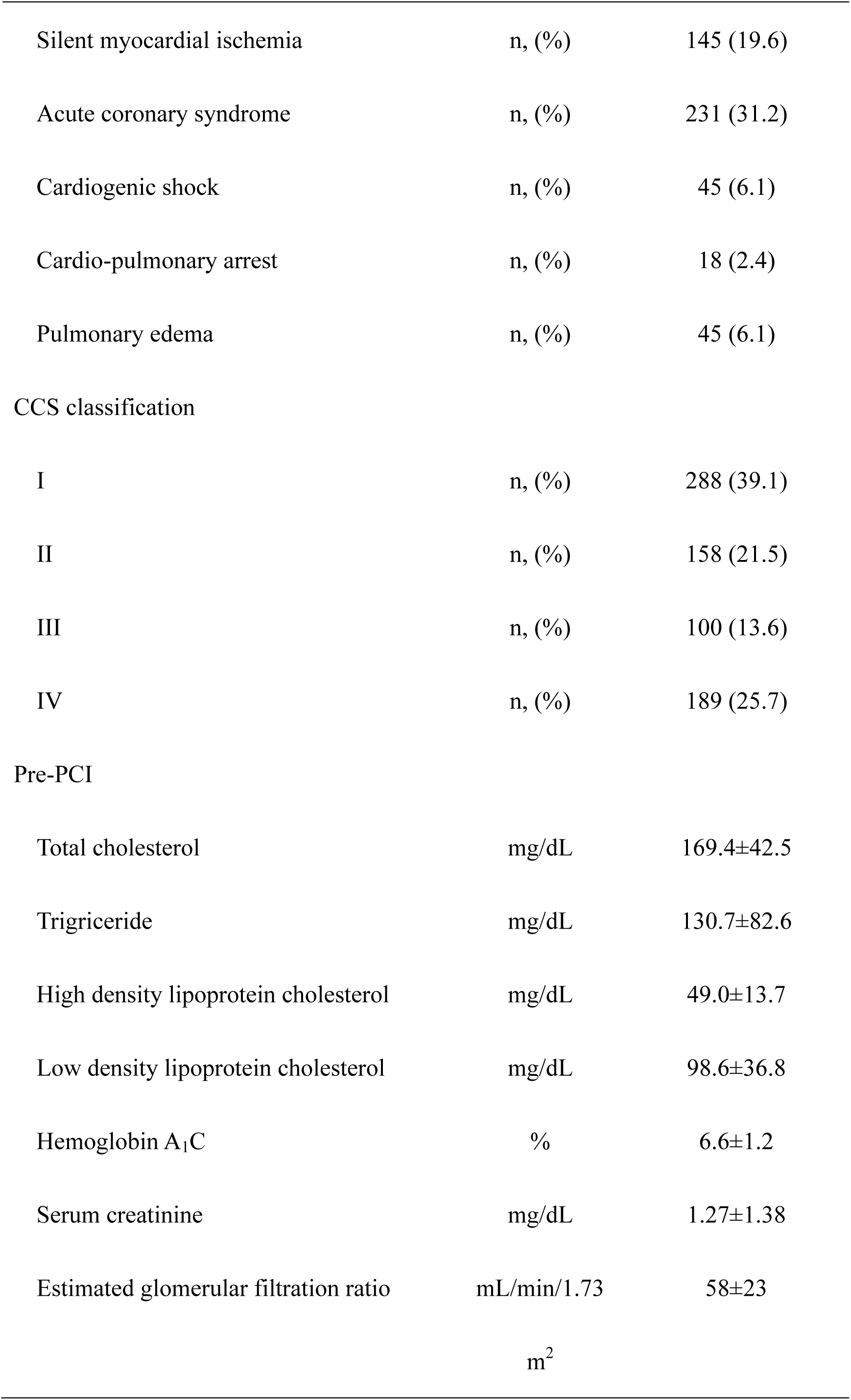
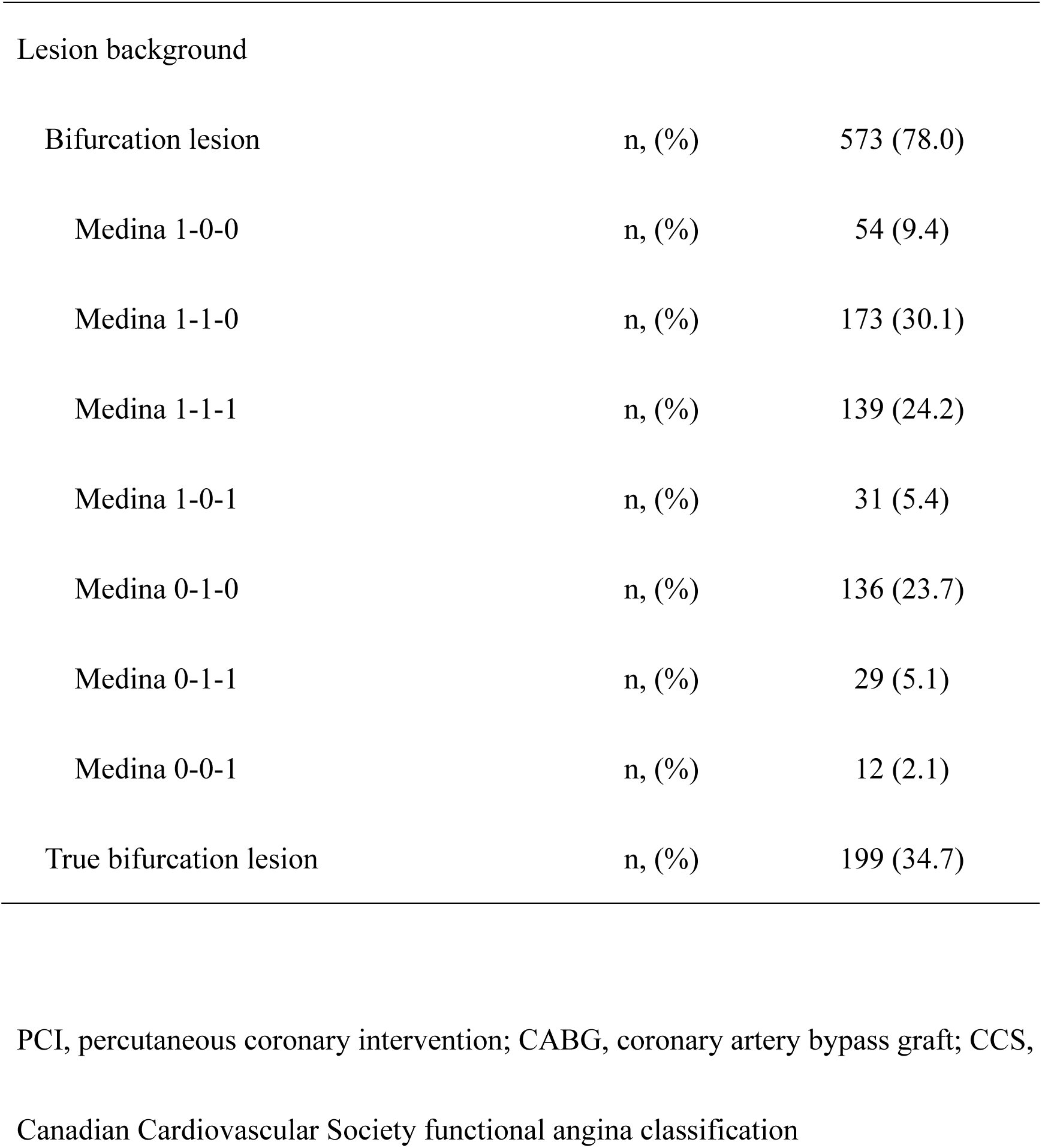
Patient and lesion background.

| Patient background |  |  |
| --- | --- | --- |
| Age | years | 73.0 ±10.1 |
| Male | n, (%) | 583 (78.7) |
| Hypertension | n, (%) | 553 (74.6) |
| Dyslipidemia | n, (%) | 526 (71.0) |
| Diabetes | n, (%) | 365 (49.3) |
| Smoking | n, (%) | 237 (32,0) |
| Hemodialysis | n, (%) | 34 (4.6) |
| Chronic obstructive pulmonary disease | n, (%) | 17 (2.3) |
| Peripheral artery disease | n, (%) | 86 (11.6) |
| Prior myocardial infarction | n, (%) | 187 (25.2) |
| Prior PCI/CABG | n, (%) | 323 (43.6) |
| Family history | n, (%) | 95 (12.8) |
| Left ventricular ejection fraction | % | 56±15 |
| Clinical presentation |  |  |
| Stable angina | n, (%) | 290 (39.2) |
| Old myocardial infarction | n, (%) | 38 (5.1) |
| Silent myocardial ischemia | n, (%) | 145 (19.6) |
| Acute coronary syndrome | n, (%) | 231 (31.2) |
| Cardiogenic shock | n, (%) | 45 (6.1) |
| Cardio-pulmonary arrest | n, (%) | 18 (2.4) |
| Pulmonary edema | n, (%) | 45 (6.1) |
| CCS classification |  |  |
| I | n, (%) | 288 (39.1) |
| II | n, (%) | 158 (21.5) |
| III | n, (%) | 100 (13.6) |
| IV | n, (%) | 189 (25.7) |
| Pre-PCI |  |  |
| Total cholesterol | mg/dL | 169.4±42.5 |
| Triglyceride | mg/dL | 130.7±82.6 |
| High density lipoprotein cholesterol | mg/dL | 49.0±13.7 |
| Low density lipoprotein cholesterol | mg/dL | 98.6±36.8 |
| Hemoglobin A<sub>1</sub>C | % | 6.6±1.2 |
| Serum creatinine | mg/dL | 1.27±1.38 |
| Estimated glomerular filtration ratio | mL/min/1.73 | 58±23 |
|  | m<sup>2</sup> |  |
| Lesion background |  |  |
| Bifurcation lesion | n, (%) | 573 (78.0) |
| Medina 1-0-0 | n, (%) | 54 (9.4) |
| Medina 1-1-0 | n, (%) | 173 (30.1) |
| Medina 1-1-1 | n, (%) | 139 (24.2) |
| Medina 1-0-1 | n, (%) | 31 (5.4) |
| Medina 0-1-0 | n, (%) | 136 (23.7) |
| Medina 0-1-1 | n, (%) | 29 (5.1) |
| Medina 0-0-1 | n, (%) | 12 (2.1) |
| True bifurcation lesion | n, (%) | 199 (34.7) |

PCI, percutaneous coronary intervention; CABG, coronary artery bypass graft; CCS,
3 Canadian Cardiovascular Society functional angina classification

### Lesion background

The backgrounds of the lesions are shown in Table 1. LM bifurcation was observed in 78.0% of the patients, and the most prevalent Medina classification was 1-1-0 lesion (30.1%) and true bifurcation lesion, which included significant stenosis in both the main vessel (MV) and SB, accounted for 34.7%. Angiographically nonsignificant LM lesions (Medina 0-x-x) were included in 30.9% of the patients because of the identification of LM atherosclerotic plaques on intracoronary imaging.

### PCI procedures

The PCI procedures are presented in Table 2. Radial access was frequently used in 70.1% of the patients, and most patients were treated with 6- and 7-Fr guiding systems (51.9% and 41.8%, respectively). Imaging guidance was routinely performed (97.7%), and IVUS and OCT/OFDI were performed in 86.5% and 12.2% of the patients, respectively. A current-generation DES was implanted in all patients, except for one in which a bare metal stent was used according to the physician’s discretion. Two-stent deployment in the LM bifurcation was performed in 8.8% of the patients, and elective procedure was performed in only 14 patients (1.9% of the entire cohort). The stenting technique was performed in the sequence of culottes, T-stenting, and crush stenting. The POT was performed in 53.1% of the patients treated with one-stent deployment. KBI or SB dilation alone was performed in 75.0% of the patients. A mechanical cardiac support device was used in 16.4% of the patients (intra-aortic balloon pumping, 15.8%; extracorporeal membrane oxygenation, 2.7%). Modification of calcified lesions was performed using rotational or orbital atherectomy (9.2%) and a scoring balloon (19.7%). MV stent size was 3.5–4.0 mm in 63.0% and 3.0–3.5 mm in 30.8% of the patients, and SB stent size was 3.0–3.5 mm in 41.0% and 2.5–3.0 mm in 34.9% of the patients. The PCI success rate was 98.6%.

**Table 2.** Procedures of left main percutaneous coronary intervention.

|  |  |  |
| --- | --- | --- |
| Access |  |  |
| Radial | n, (%) | 519 (70.1) |
| Femoral | n, (%) | 198 (26.8) |
| Brachial | n, (%) | 23 (3.1) |
| System |  |  |
| 6 Fr | n, (%) | 383 (51.9) |
| 7 Fr | n, (%) | 309 (41.8) |
| 8 Fr | n, (%) | 47 (6.4) |
| Imaging guide | n, (%) | 726 (97.7) |
| Optical coherence tomography | n, (%) | 90 (12.2) |
| Drug-eluting stent |  |  |
| First generation | n, (%) | 0 (0) |
| Current generation | n, (%) | 742 (99.9) |
| Two-stent implantation | n, (%) | 65 (8.8) |
| Elective | n, (%) | 14 (21.5) |
| Culotte | n, (%) | 26 (40.0) |
| Crush | n, (%) | 16 (24.6) |
| T-stenting | n, (%) | 21 (32.3) |
| Proximal optimization technique | n, (%) | 333 (53.1) |
| Side branch dilation in 1-stent | n, (%) | 472 (75.0) |
| Kissing balloon inflation | n, (%) | 390 (62.0) |
| Side branch dilation alone | n, (%) | 82 (13.0) |
| Mechanical cardiac support device | n, (%) | 122 (16.4) |
| Intra-aortic balloon pumping | n, (%) | 117 (15.8) |
| Extracorporeal membrane oxygenation | n, (%) | 20 (2.7) |
| Lesion modification |  |  |
| Rotational/Orbital atherectomy | n, (%) | 68 (9.2) |
| Directional coronary atherectomy | n, (%) | 10 (1.4) |
| Scoring balloon | n, (%) | 145 (19.7) |
| Main vessel stent |  |  |
| Size: $\geq 4.0$ mm | n, (%) | 117 (16.0) |
| 3.5–4.0 mm | n, (%) | 390 (53.3) |
| 3.0–3.5 mm | n, (%) | 187 (25.5) |
| 2.5–3.0 mm | n, (%) | 33 (4.5) |
| <2.5 mm | n, (%) | 5 (0.7) |
| Length | mm | 23.0±8.5 |
| Product |  |  |
| Xience | n, (%) | 359 (49.4) |
| Synergy/Promus | n, (%) | 129 (17.8) |
| Resolute Integrity/Onyx | n, (%) | 79 (10.9) |
| Ultimaster | n, (%) | 106 (14.6) |
| Nobori/BioFreedom | n, (%) | 38 (5.2) |
| Orsilo | n, (%) | 14 (1.9) |
| Bare metal stent | n, (%) | 1 (0.1) |
| Side branch |  |  |
| Size: ≥4.0 mm | n, (%) | 4 (4.8) |
| 3.5–4.0 mm | n, (%) | 7 (8.4) |
| 3.0–3.5 mm | n, (%) | 34 (41.0) |
| 2.5–3.0 mm | n, (%) | 29 (34.9) |
| <2.5 mm | n, (%) | 9 (10.8) |
| Length | mm | 21.6±7.8 |
| Product |  |  |
| Xience | n, (%) | 34 (41.0) |
| Synergy/Promus | n, (%) | 16 (19.3) |
| Resolute Integrity/Onyx | n, (%) | 13 (15.7) |
| Ultimaster | n, (%) | 17 (20.5) |
| Nobori/BioFreedom | n, (%) | 1 (1.2) |
| Orsilo | n, (%) | 2 (2.4) |
| Bare metal stent | n, (%) | 0 (0) |
| Procedural success | n, (%) | 730 (98.6) |

### Adverse events at 1-year follow-up

Adverse events at the 1-year follow-up are shown in Table 3. The rates of all-cause death, cerebrovascular disorders, clinically driven revascularization, and myocardial infarction were 8.9%, 1.2%, 8.2%, and 1.9%, respectively. MACCE, which were a composite of these events, were observed in 17.5% of the patients. TLR and cardiac death occurred in 2.0% and 3.4% of the patients, respectively. The cumulative curve of MACCE showed a gradual increase (Figure 2a), whereas that of all-cause death peaked within 30 days and plateaued at 1 year (Figure 3a).

**Figure 2.**
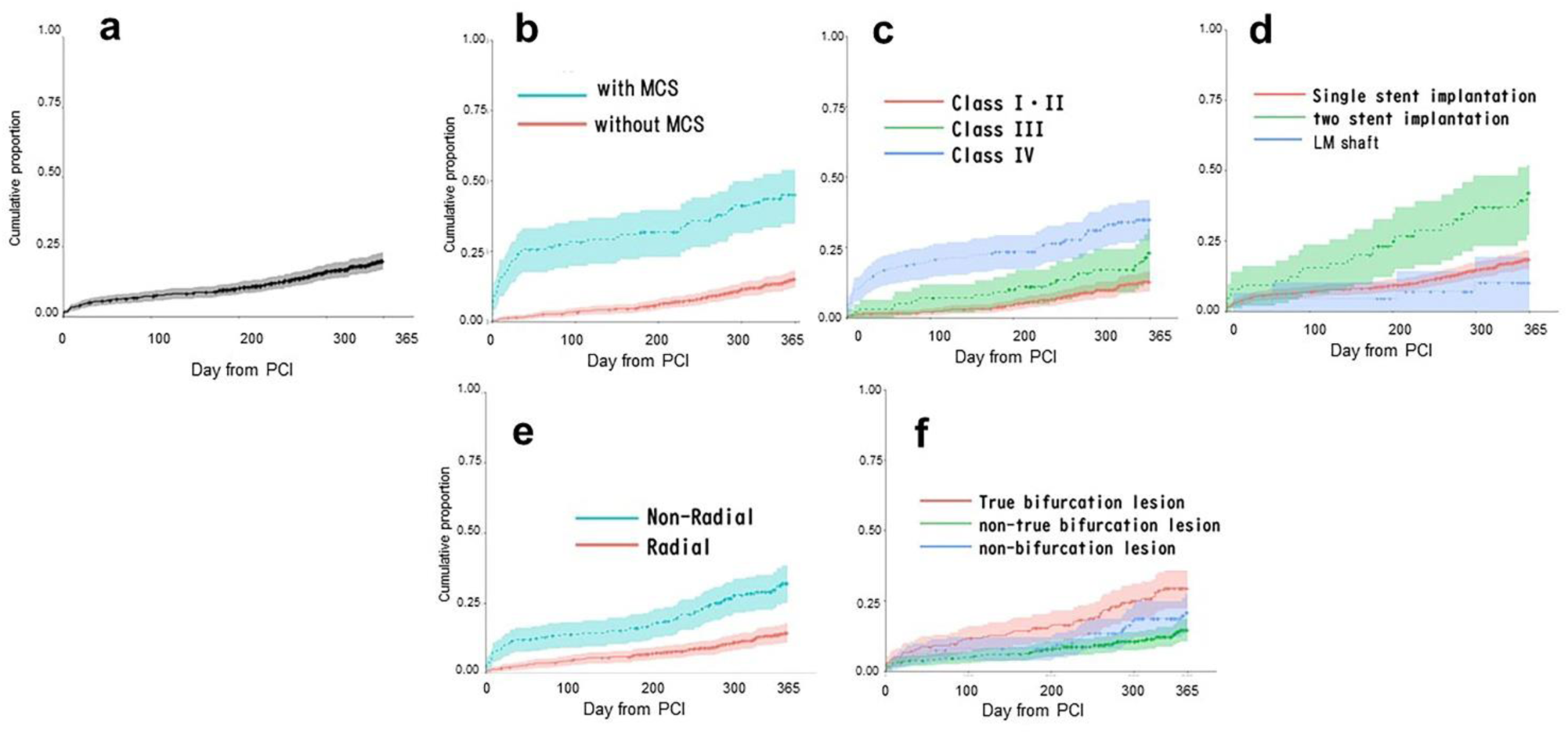
Cumulative curve of major adverse cardiovascular and cerebrovascular events. (a) Entire group. (b) The groups using or not using a mechanical cardiac support device (MCS). (c) Canadian Cardiovascular Society functional angina classification. (d) The groups treated with a single stent, two stents, and left main shaft stenting alone. (e) The groups of radial and nonradial access. (f) The groups of true bifurcation lesion, non-true bifurcation lesion, and non-bifurcation lesion.

**Figure 3.**
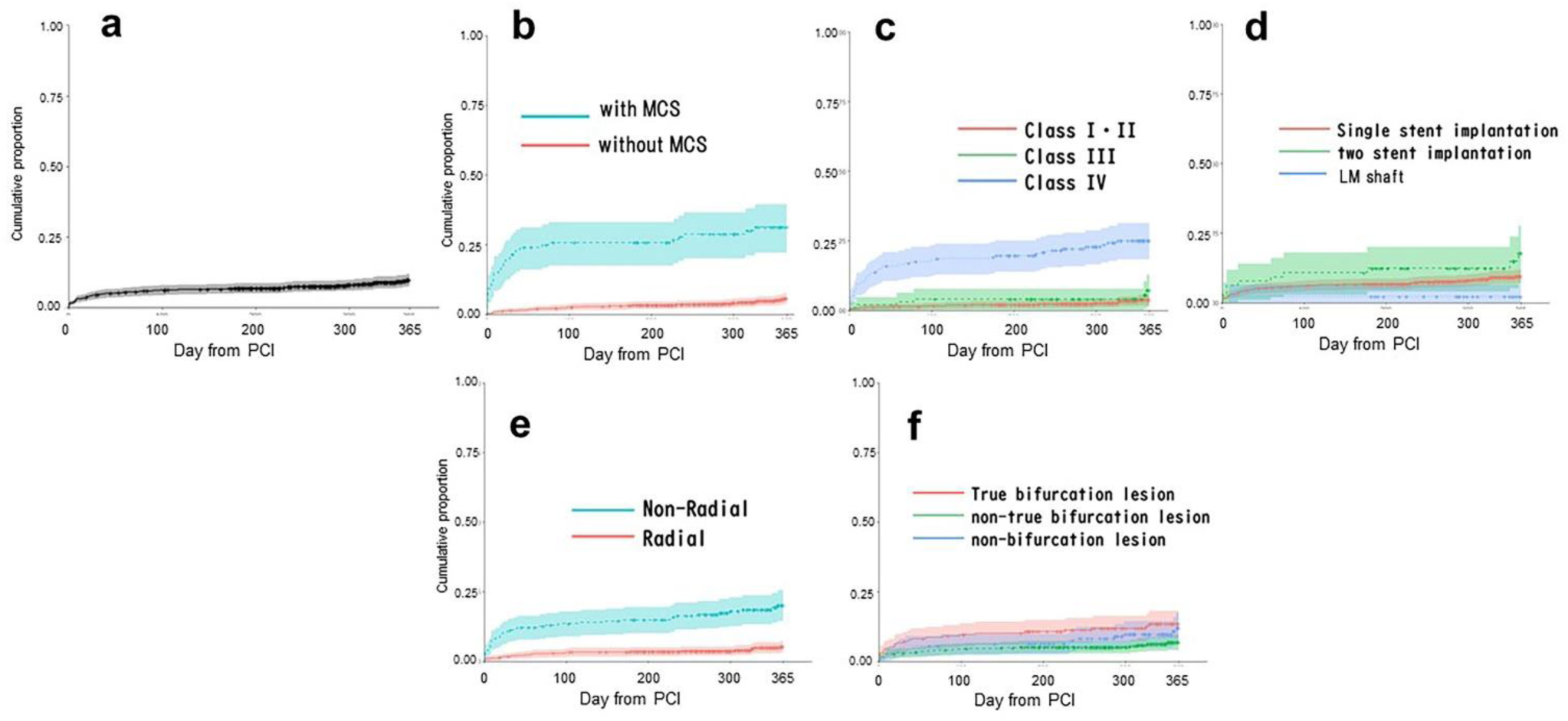
Cumulative curve of all-cause mortality. (a) Entire group. (b) The groups using or not using a mechanical cardiac support device (MCS). (c) Canadian Cardiovascular Society functional angina classification. (d) The groups treated with a single stent, two stents, and left main shaft stenting alone. (e) The groups of radial and nonradial access. (f) The groups of true bifurcation lesion, non-true bifurcation lesion, and non-bifurcation lesion.

**Table 3.**
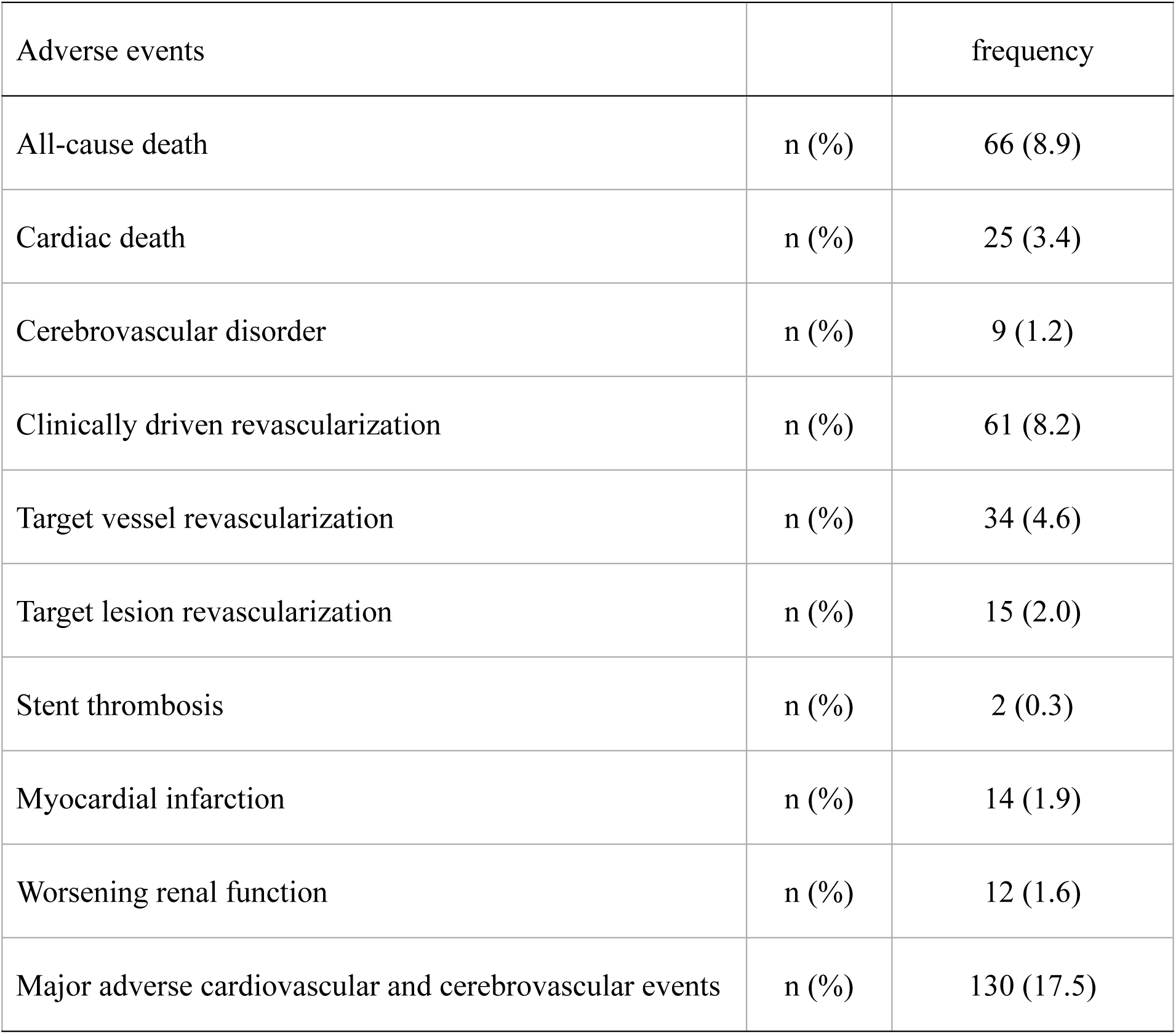
Clinical outcomes at the 1-year follow-up.

| Adverse events |  | frequency |
| --- | --- | --- |
| All-cause death | n (%) | 66 (8.9) |
| Cardiac death | n (%) | 25 (3.4) |
| Cerebrovascular disorder | n (%) | 9 (1.2) |
| Clinically driven revascularization | n (%) | 61 (8.2) |
| Target vessel revascularization | n (%) | 34 (4.6) |
| Target lesion revascularization | n (%) | 15 (2.0) |
| Stent thrombosis | n (%) | 2 (0.3) |
| Myocardial infarction | n (%) | 14 (1.9) |
| Worsening renal function | n (%) | 12 (1.6) |
| Major adverse cardiovascular and cerebrovascular events | n (%) | 130 (17.5) |

### Risk in major adverse cardiovascular and cerebrovascular events

Multivariate analysis of the risk of MACCE is shown in Table 4. Independent factors that elevated the risk included CCS ≥class III (HR, 2.07; 95% CI, 1.39–3.07; p<0.001), mechanical cardiac support device use (HR, 2.17; 95% CI, 1.43–3.30; p<0.001), and two-stent implantation (HR, 2.49; 95% CI, 1.48–4.18; p<0.001). Factors that reduced the risk were left ventricular ejection fraction −10% increase (HR, 0.72; 95% CI, 0.52–0.99; p=0.041) and radial access (HR, 0.62; 95% CI, 0.42–0.91; p=0.015). The cumulative curve of MACCE showed higher rates of mechanical cardiac support device use (Figure 2b), CCS classes III and IV (c), two-stent implantation (d), and nonradial access (e). The cumulative curve of all-cause death showed higher rates of mechanical cardiac support device use (Figure 3b), CCS class IV (c), and nonradial access (e), which peaked within 30 days.

**Table 4.** Multivariate analysis of the risk of major adverse cardiovascular and cerebrovascular events.

| Variables | Hazard ratio | 95% CI | P value |
| --- | --- | --- | --- |
| Age (10 years increase) | 1.00 | 0.79–1.27 | 0.979 |
| Male | 1.13 | 0.71–1.81 | 0.597 |
| Smoker | 1.31 | 0.89–1.93 | 0.178 |
| Body mass index (1 kg/m<sup>2</sup> increase) | 1.04 | 0.95–1.14 | 0.372 |
| eGFR (10 mL/min/1.73 m<sup>2</sup> increase) | 0.84 | 0.67–1.04 | 0.107 |
| Left ventricular ejection fraction (10% increase) | 0.72 | 0.52–0.99 | 0.041 |
| Dyslipidemia | 0.90 | 0.60–1.33 | 0.592 |
| Hypertension | 1.19 | 0.76–1.86 | 0.438 |
| Diabetes mellitus | 0.99 | 0.68–1.43 | 0.952 |
| CCS ≥class III | 2.07 | 1.39–3.07 | <0.001 |
| Residual significant stenosis after PCI | 1.31 | 0.85–2.00 | 0.221 |
| Radial access | 0.62 | 0.42–0.91 | 0.015 |
| Mechanical cardiac support device | 2.17 | 1.43–3.30 | <0.001 |
| Lesion modification: rotational or orbital | 1.40 | 0.78–2.53 | 0.260 |
| atherectomy |  |  |  |
| Left main stent size >3.5 mm | 0.75 | 0.51–1.10 | 0.140 |
| Two-stent implantation | 2.49 | 1.48–4.18 | 0.001 |
| True bifurcation lesion | 0.91 | 0.55–1.48 | 0.693 |
| Proximal optimization technique | 0.89 | 0.62–1.29 | 0.542 |

CI, confidence interval; eGFR, estimated glomerular filtration ratio; CCS, Canadian
Cardiovascular Society functional angina classification

## Discussion

In the LM-JANHO study on routine imaging-guided unprotected LM-PCI, including acute coronary syndrome, 17.5% of the patients experienced composite endpoints (comprising all-cause death, cerebrovascular disorder, any clinically driven revascularization, and myocardial infarction) at the 1-year follow-up. Notably, the study reported a low incidence of TLR (2.0%) and cardiac death (3.4%). Risk factors for the composite endpoints included CCS class ≥III, use of a mechanical cardiac support device, and two-stent implantation, while a 10% increase in left ventricular ejection fraction and radial access were associated with a reduced risk.

### Effect of routine imaging guidance in LM-PCI

Imaging guidance in LM-PCI demonstrated a lower incidence of composite endpoints of all-cause death, restenosis, or definite stent thrombosis (HR, 0.65; 95% CI, 0.50–0.84) and all-cause death alone (HR, 0.62; 95% CI, 0.47–0.82) in 2468 patients of the Swedish Coronary Angiography and Angioplasty Registry, in which IVUS guidance was used in 25.2% of the patients^2^. In 11,264 unprotected LM-PCI procedures performed in England and Wales, analysis of 5056 pairs of imaging- and angio-guided PCIs after propensity matching showed that the imaging-guided PCI presented a lower incidence of 12-month mortality than the angio-guided PCI (odds ratio [OR], 0.660; 95% CI, 0.57–0.77)^7^. A meta-analysis of comparison between IVUS guidance (2096 patients) and angio-guidance (1984 patients) in LM-PCI also exhibited a lower incidence of all-cause death (OR, 0.46; 95% CI, 0.30–0.71), cardiac death (OR, 0.47; 95% CI, 0.29–0.77), and composite endpoints, including stent thrombosis, myocardial infarction, cardiac death, and all-cause death (OR, 0.42; 95% CI, 0.26–0.67)^3^. However, imaging guidance was employed for complex lesions in these studies, and the efficacy of routine imaging in every LM-PCI has not been elucidated.

The present study included cases with acute myocardial infarction and cardiogenic shock that were excluded in the other previous studies of the imaging-guided LM-PCI^3, 5–7, 17^. Therefore, all-cause death (8.9% vs. 2.5–9.0%) and cardiac death at 1 year (3.4% vs. 0.8–2.4%) were observed more frequently in the present study than in previous studies. However, there was a similar incidence rate of TVR (4.6% vs. 4.2–6.7%) and lower incidence rates of TLR (2.0% vs. 2.7–7.7%), stent thrombosis (0.3% vs. 0.5–1.7%), and myocardial infarction (1.9% vs. 1.5–11.2%), indicating that optimal PCI was induced by imaging guidance in survivors after acute coronary events. The following factors contribute to the efficacy of routine imaging guidance in LM-PCI: first, the optimal device selection in terms of size and length was based on pre-PCI imaging, which decreased additional stenting due to overdilation or inappropriate positioning of the balloon or stent in the plaque-rich area. Information concerning plaque morphology, thrombus burden, and intracoronary calcification severity was also useful for appropriate preparation before stenting. Second, some atherosclerotic plaques were identified by intracoronary imaging in the angiographically insignificant LM lesions, which increased LM crossover stenting in the Medina (0-x-x) lesions. In this study, this type of lesion was observed in 30.9% of the patients, whereas in previous studies with limited use of imaging, it was observed in 0–21.5% of the patients^18–21^. Third, more appropriate PCI optimization for sufficient stent expansion, less stent malapposition or deformation, and navigation of optimal SB guidewire crossings were promoted. Accurate assessment of SB dissection or stenosis after balloon dilation on intracoronary imaging resulted in a decrease in two-stent deployment in this study (8.8%). Elective two-stent deployment was performed in only 1.9% of the patients, and fewer escalations from provisional stenting (6.9%) were noted. Thus, local effectiveness of imaging-guided PCI on LM lesion was prominent in less TLR (2.0%) and cardiac death (3.4%) in this study compared with the prior Japanese registry study (AOI registry) (TLR: 9.1% [bifurcation]/11.0% [non-bifurcation], cardiac death: 6.6%/5.0%^19^) or randomized studies comparing provisional and elective two stenting with the limited use of imaging guidance and exclusion of acute myocardial infarction and cardiogenic shock (EBC MAIN, TLR: 6.1% [provisional stent]/9.3% [two stents]^21^; DK Crush V, TLR: 10.7%/5.0%, cardiac death: 1.3%/2.1%^20^).

### Risk of MACCE

CCS class ≥III and mechanical cardiac support device use were directly related to the severe hemodynamic collapse state, which might result in cardiac death in acute phase. As nonradial access was adopted more frequently in an unstable hemodynamic state with weak pulsation of the radial artery, it was also associated with the risk of cardiac death. In fact, the group with these factors had elevated mortality within 30 days and reached a plateau at 1 year (Figure 3). Even with technical improvements in LM-PCI under imaging guidance, the initial myocardial damage induced by LM ischemia cannot be effectively relieved.

Lower left ventricular ejection fraction was identified as an independent risk factor for MACCE. This was a result of LM ACS that led to severe left ventricular damage or ischemic cardiomyopathy that had already presented with multiple-vessel diseases, including LM lesions. A lower left ventricular ejection fraction was associated with heart failure recurrence, fatal arrhythmic events, and more frequent coronary revascularization.

Two-stent deployment was also an independent risk factor for MACCE. Higher risk^18, 19, 22, 23^ of two-stent deployment was demonstrated in previous studies in which first-generation DES were mainly used. However, similar^21, 23^ or superior clinical outcomes^20, 24^ compared with provisional stenting have been reported in recent studies that used current-generation DES. In the present study, two-stent deployment was used in a limited manner for more complex lesions (8.8%). More than 20% of the cases that would have been treated with elective two-stent deployment in other studies with limited use of intracoronary imaging were treated with provisional stenting in this study. However, true bifurcation lesions were not identified as risk factors for MACCE, and some SB lesions were left with dissection or incomplete full vessel dilation. Angiographically hazy images after pre-dilation in the SB were likely to promote more elective two-stent deployment or escalation to provisional two stenting in other studies. Escalation to provisional two stenting was observed in 22–47% of the patients owing to suboptimal findings in the SB, which were mainly assessed by angiographical diameter stenosis^25^ and have been revealed to be overestimated with physiologically insignificant stenosis in 30% of the patients^26^. Although definite imaging criteria were not established in this study, individual operators decided to perform additional SB stenting more strictly according to imaging findings, such as intimal flap, which threatens vessel occlusion, progression of hematoma, and insufficient lumen dilation with concrete plaque remaining. Although suboptimal angiographic findings remained in some true bifurcation lesions, they were not associated with worse clinical outcomes in imaging-guided LM-PCI. As most two-stent deployments were adopted in cases with suboptimal SB balloon dilation or multiple vessel involvement, diffuse lesions, hard calcification, rich plaque burden, and stent landing in the distal plaque were likely to be present, requiring more vessel revascularization. Therefore, comprehensive management strategies that consider plaque debulking, aggressive medical therapy, and switching to CABG after lesion assessment using intracoronary imaging are required.

### Contemporary PCI techniques

#### Radial access

In the present study, radial access was used in 70.1% of the patients using a 6- or 7-Fr guiding catheter. Radial access was first approached in elective cases, and the introduction of a slender system with a thinner outer sheath by 1 Fr than the conventional system allowed for increased use of a 7-Fr guiding catheter, which enabled complex PCI procedures to be performed with greater ease. Radial access is associated with less bleeding (OR, 0.43; 95% CI, 0.27–0.69; p=0.0004) and lower in-hospital mortality rate (OR, 0.49; 95% CI, 0.31–0.79; p=0.004) in a meta-analysis of LM-PCI including 17,258 patients^27^.

#### POT

In the present study, POT was performed in 53.1% of the patients, which was not as in randomized trials^8, 20, 21^, but more frequent than in other registry studies^6, 19, 28^. The POT provides sufficient stent expansion and less stent malapposition in the proximal MV, and re-POT is recommended after SB dilation to correct stent deformation^28–30^. In the present study, the more frequent KBI (62%) might have resulted in sufficient stent expansion with less malapposition in the LM, and confirmation by imaging led to a lower performance of POT or re-POT. However, it is crucial to determine the optimal size of the balloon for the POT according to the LM vessel size in the pre-PCI imaging and to confirm sufficient stent expansion in the LM, including the polygon confluence without any significant stent malapposition in the post-PCI imaging^31^.

#### Optimal SB wiring

In imaging-guided bifurcation PCI, confirmation of the optimal SB wiring before SB dilation is recommended^11^. Even in two-dimensional IVUS imaging, the optimal SB guidewire position can be identified as the most distal site at the carina between the struts located at the center of the carinal site of the SB. Three-dimensional reconstruction of OCT/OFDI images provides a clear image of the SB guidewire recrossing point and the configuration of the SB jailing strut pattern^32, 33^. Three-dimensional OCT/OFDI guidance enhances optimal SB wiring to the distal cell at the SB ostium (from 50–66% to 90%) and reduces incomplete stent apposition after subsequent KBI^32, 34^. In the present study, OCT/OFDI guidance was infrequent (12.2%); however, confirmation of the SB wiring position was routinely performed under IVUS guidance.

#### SB dilation under the imaging guidance

Any SB dilation was performed more frequently (75%) in the present study than in other registry studies^26, 28, 35, 36^. The LCX ostial area is larger than that of other SBs in non-LM bifurcations and is covered with more jailing struts^32^. The success rate of optimal SB wiring was lower in LM bifurcation than in non-LM bifurcation under angio-guidance (55% vs. 75%), and more incomplete strut apposition was observed in LM bifurcation^32^. Incomplete removal of jailing struts in the LCX ostium due to suboptimal SB wiring or inappropriate balloon dilation leads to clustering of jailing struts at the rim of the LCX ostium, which may be associated with thrombus attachment or restenotic reaction^37–39^. More suboptimal SB wiring and subsequent SB dilation than expected would have occurred in previous studies due to the limited use of imaging. The efficacy of KBI in angiographic SB restenosis and the reduction of subclinical thrombus attachment to the jailing struts was confirmed in a study of imaging-guided bifurcation PCI^32, 40^. Although leaving the LCX without any dilation after LM-LAD crossover stenting has a sufficiently higher survival rate from cardiac events in cases with high FFR values in the jailed LCX^35^, there are reports of non-fenestration-related restenosis due to intimal coverage of the jailing struts at the LCX ostium and very late stent thrombosis on the jailing struts in an autopsy study of sudden cardiac death of patients after LM-PCI^39^. In the e-Ultimaster bifurcation substudy, which demonstrated the efficacy of the POT but not that of KBI, the least and most prevalent target lesion failures were found in cases with both POT and KBI performed and those without either of them, which indicates that leaving the SB untouched without any optimization in the proximal MV or SB is associated with worse clinical outcomes^28^. Imaging-guided optimal SB treatment and POT are crucial to reduce procedure-related events.

#### Mechanical cardiac supporting device

In the present study, intra-aortic balloon pumping was used in 15.8% of the patients, and extracorporeal membrane oxygenation was used in 2.7% of the patients. As Impella had not been introduced in Japan at the start of the study period and had limited use for cardiogenic shock in licensed institutes, there were no cases with Impella support in this study. As the inefficacy of intra-aortic balloon pumping or extracorporeal membrane oxygenation on long-term survival has been reported^41, 42^ a higher rate of cardiac death in patients using a mechanical cardiac support device was demonstrated in this study. Improvement of the survival rate after LM-related ACS has been reported in Impella support due to early unloading of the left ventricle, prevention of its remodeling, and subsequent multiple organ failure^43^. Improvement in mortality is expected using Impella support; however, further studies are warranted to investigate its efficacy in LM-related ACS in East Asian races, which have shown a higher bleeding risk than Caucasian races^44^.

### Study limitations

The present study has the following limitations. 1) The study did not adopt a randomized trial design, with 97.7% of cases treated under imaging guidance. Consequently, the study design does not facilitate a direct assessment of the efficacy of imaging guidance in comparison with angio-guidance. Instead, efficacy was established by comparing the results with those from previous reports. 2) Given that consecutive cases of LM-PCI were enrolled, this study included cases of acute myocardial infarction or cardiogenic shock, which were excluded in previous studies. This may have led to an increase in clinical events, particularly in terms of mortality. 3) Because this was a retrospective study, there were no standardized criteria for optimal PCI results in intracoronary imaging among the participating institutes, except for the general recommendations of a stent expansion index >80%, minimal stent malapposition, and confirmation of the SB wiring site. 4) The enrollment of LM-PCI or CABG in LM-related diseases varied among the participating institutes, and the expertise and skills of operators performing LM-PCI also differed. These variations may have influenced the study outcomes.

## Data Availability

The data that support the findings of this study are available from the corresponding author, [Yoshinobu Murasato], upon reasonable request.

## Non-standard Abbreviations and Acronyms

CABG: coronary artery bypass graft
CCS: Canadian Cardiovascular Society functional angina classification
CI: confidence interval
DES: drug-eluting stent
HR: hazard ratio
IVUS: intravascular ultrasound
KBI: kissing balloon inflation
LAD: left anterior descending artery
LCX: left circumflex artery
LM: left main
LM-JANHO: left main coronary intervention in a cohort of Japanese National Hospital Organization
MACCE: major adverse cardiovascular and cerebrovascular events
MV: main vessel
OFDI: optical frequency dependent imaging
OCT: optical coherence tomography
OR: odds ratio
PCI: percutaneous coronary intervention
POT: proximal optimal technique
SB: side branch
SYNTAX: Synergy between Percutaneous Coronary Intervention with Taxus and Cardiac Surgery
TLR: target lesion revascularization
TVR: target vessel revascularization

## Acknowledgements

None.

## References

1. Buccheri S, Franchina G, Romano S, Puglisi S, Venuti G, D’Arrigo P, Francaviglia B, Scalia M, Condorelli A, Barbanti M, et al. Clinical Outcomes Following Intravascular Imaging-Guided Versus Coronary Angiography-Guided Percutaneous Coronary Intervention With Stent Implantation: A Systematic Review and Bayesian Network Meta-Analysis of 31 Studies and 17,882 Patients. JACC Cardiovasc Interv. 2017;10:2488–2498.

2. Andell P, Karlsson S, Mohammad MA, Gotberg M, James S, Jensen J, Frobert O, Angeras O, Nilsson J, Omerovic E, et al. Intravascular Ultrasound Guidance Is Associated With Better Outcome in Patients Undergoing Unprotected Left Main Coronary Artery Stenting Compared With Angiography Guidance Alone. Circ Cardiovasc Interv. 2017;10:e004813.

3. Zhang Q, Wang B, Han Y, Sun S, Lv R, Wei S. Short- and Long-Term Prognosis of Intravascular Ultrasound-Versus Angiography-Guided Percutaneous Coronary Intervention: A Meta-Analysis Involving 24,783 Patients. J Interv Cardiol. 2021;2021:6082581.

4. Lee JM, Choi KH, Song YB, Lee JY, Lee SJ, Lee SY, Kim SM, Yun KH, Cho JY, Kim CJ, et al. Intravascular Imaging-Guided or Angiography-Guided Complex PCI. N Engl J Med. 2023;388:1668–1679.

5. de la Torre Hernandez JM, Garcia Camarero T, Baz Alonso JA, Gomez-Hospital JA, Veiga Fernandez G, Lee Hwang DH, Sainz Laso F, Sanchez-Recalde A, Perez de Prado A, Lozano Martinez-Luengas I, et al. Outcomes of Predefined Optimisation Criteria for Intravascular Ultrasound Guidance of Left Main Stenting. EuroIntervention. 2020;16:210–217.

6. Cortese B, Piraino D, Gentile D, Onea HL, Lazar L. Intravascular Imaging for Left Main Stem Assessment: An Update on the Most Recent Clinical Data. Catheter Cardiovasc Interv. 2022;100:1220–1228.

7. Kinnaird T, Johnson T, Anderson R, Gallagher S, Sirker A, Ludman P, de Belder M, Copt S, Oldroyd K, Banning A, et al. Intravascular Imaging and 12-Month Mortality After Unprotected Left Main Stem PCI: An Analysis From the British Cardiovascular Intervention Society Database. JACC Cardiovasc Interv. 2020;13:346–357.

8. Holm NR, Andreasen LN, Neghabat O, Laanmets P, Kumsars I, Bennett J, Olsen NT, Odenstedt J, Hoffmann P, Dens J, et al. OCT or Angiography Guidance for PCI in Complex Bifurcation Lesions. N Engl J Med. 2023;389:1477–1487.

9. Neumann FJ, Sousa-Uva M, Ahlsson A, Alfonso F, Banning AP, Benedetto U, Byrne RA, Collet JP, Falk V, Head SJ, et al. 2018 ESC/EACTS Guidelines on Myocardial Revascularization. Eur Heart J. 2019;40:87–165.

10. Nakamura M, Yaku H, Ako J, Arai H, Asai T, Chikamori T, Daida H, Doi K, Fukui T, Ito T, et al. JCS/JSCVS 2018 Guideline on Revascularization of Stable Coronary Artery Disease. Circ J. 2022;86:477–588.

11. Takagi K, Nagoshi R, Kim BK, Kim W, Kinoshita Y, Shite J, Hikichi Y, Song YB, Nam CW, Koo BK, et al. Efficacy of Coronary Imaging on Bifurcation Intervention. Cardiovasc Interv Ther. 2021;36:54–66.

12. Raber L, Mintz GS, Koskinas KC, Johnson TW, Holm NR, Onuma Y, Radu MD, Joner M, Yu B, Jia H, et al. Clinical Use of Intracoronary Imaging. Part 1: Guidance and Optimization of Coronary Interventions. An Expert Consensus Document of the European Association of Percutaneous Cardiovascular Interventions. Eur Heart J. 2018;39:3281–3300.

13. Murasato Y, Kinoshita Y, Yamawaki M, Okamura T, Nagoshi R, Watanabe Y, Suzuki N, Mori T, Shinke T, Shite J, et al. Impact of Medina Classification on Clinical Outcomes of Imaging-guided Coronary Bifurcation Stenting. Int J Cardiol Heart Vasc. 2023;49:101311.

14. Windecker S, Neumann FJ, Juni P, Sousa-Uva M, Falk V. Considerations for the Choice Between Coronary Artery Bypass Grafting and Percutaneous Coronary Intervention as Revascularization Strategies in Major Categories of Patients with Stable Multivessel Coronary Artery Disease: An Accompanying Article of the Task force of the 2018 ESC/EACTS Guidelines on Myocardial Revascularization. Eur Heart J. 2019;40:204–212.

15. Kameyama T, Ino Y, Kubo T, Akasaka T. Advances in Coronay Artery Imaging (in Japanese). J Jpn Coron Assoc. 2016;22:39–44.

16. Sawano M, Yamaji K, Kohsaka S, Inohara T, Numasawa Y, Ando H, Iida O, Shinke T, Ishii H, Amano T. Contemporary Use and Trends in Percutaneous Coronary Intervention in Japan: An Outline of the J-PCI Registry. Cardiovasc Interv Ther. 2020;35:218–226.

17. Maznyczka A, Arunothayaraj S, Egred M, Banning A, Brunel P, Ferenc M, Hovasse T, Wlodarczak A, Pan M, Schmitz T, et al. Bifurcation Left Main Stenting With or Without Intracoronary Imaging: Outcomes from the EBC MAIN Trial. Catheter Cardiovasc Interv. 2023;102:415–429.

18. Toyofuku M, Kimura T, Morimoto T, Hayashi Y, Shiode N, Nishikawa H, Nakao K, Shirota K, Kawai K, Hiasa Y, et al. Comparison of 5-year Outcomes in Patients With and Without Unprotected Left Main Coronary Artery Disease After Treatment With Sirolimus-eluting Stents: Insights from the j-Cypher Registry. JACC Cardiovasc Interv. 2013;6:654–663.

19. Ohya M, Kadota K, Toyofuku M, Morimoto T, Higami H, Fuku Y, Yamaji K, Muranishi H, Yamaji Y, Nishida K, et al. Long-Term Outcomes After Stent Implantation for Left Main Coronary Artery (from the Multicenter Assessing Optimal Percutaneous Coronary Intervention for Left Main Coronary Artery Stenting Registry). Am J Cardiol. 2017;119:355–364.

20. Chen SL, Zhang JJ, Han Y, Kan J, Chen L, Qiu C, Jiang T, Tao L, Zeng H, Li L, et al. Double Kissing Crush Versus Provisional Stenting for Left Main Distal Bifurcation Lesions: DKCRUSH-V Randomized Trial. J Am Coll Cardiol. 2017;70:2605–2617.

21. Hildick-Smith D, Egred M, Banning A, Brunel P, Ferenc M, Hovasse T, Wlodarczak A, Pan M, Schmitz T, Silvestri M, et al. The European Bifurcation Club Left Main Coronary Stent Study: A Randomized Comparison of Stepwise Provisional vs. Systematic Dual Stenting Strategies (EBC MAIN). Eur Heart J. 2021;42:3829–3839.

22. Behan MW, Holm NR, de Belder AJ, Cockburn J, Erglis A, Curzen NP, Niemela M, Oldroyd KG, Kervinen K, Kumsars I, et al. Coronary Bifurcation Lesions Treated with Simple or Complex Stenting: 5-year Survival from Patient-level Pooled Analysis of the Nordic Bifurcation Study and the British Bifurcation Coronary Study. Eur Heart J. 2016;37:1923–1928.

23. Lee JM, Hahn JY, Kang J, Park KW, Chun WJ, Rha SW, Yu CW, Jeong JO, Jeong MH, Yoon JH, et al. Differential Prognostic Effect Between First- and Second-Generation Drug-Eluting Stents in Coronary Bifurcation Lesions: Patient-Level Analysis of the Korean Bifurcation Pooled Cohorts. JACC Cardiovasc Interv. 2015;8:1318–1331.

24. Zhang JJ, Ye F, Xu K, Kan J, Tao L, Santoso T, Munawar M, Tresukosol D, Li L, Sheiban I, et al. Multicentre, randomized comparison of two-stent and provisional stenting techniques in patients with complex coronary bifurcation lesions: the DEFINITION II trial. Eur Heart J. 2020;41:2523–2536.

25. Paradies V, Banning A, Cao D, Chieffo A, Daemen J, Diletti R, Hildick-Smith D, Kandzari DE, Kirtane AJ, Mehran R, et al. Provisional Strategy for Left Main Stem Bifurcation Disease: A State-of-the-Art Review of Technique and Outcomes. JACC Cardiovasc Interv. 2023;16:743–758.

26. Lee HS, Kim U, Yang S, Murasato Y, Louvard Y, Song YB, Kubo T, Johnson TW, Hong SJ, Omori H, et al. Physiological Approach for Coronary Artery Bifurcation Disease: Position Statement by Korean, Japanese, and European Bifurcation Clubs. JACC Cardiovasc Interv. 2022;15:1297–1309.

27. Goel S, Pasam RT, Raheja H, Gotesman J, Gidwani U, Ahuja KR, Reed G, Puri R, Khatri JK, Kapadia SR. Left Main Percutaneous Coronary Intervention-Radial Versus Femoral Access: A Systematic Analysis. Catheter Cardiovasc Interv. 2020;95:E201–E213.

28. Chevalier B, Mamas MA, Hovasse T, Rashid M, Gomez-Hospital JA, Pan M, Witkowski A, Crowley J, Aminian A, McDonald J, et al. Clinical Outcomes of the Proximal Optimisation Technique (POT) in Bifurcation Stenting. EuroIntervention. 2021;17:e910–e918.

29. Albiero R, Burzotta F, Lassen JF, Lefevre T, Banning AP, Chatzizisis YS, Johnson TW, Ferenc M, Pan M, Daremont O, et al. Treatment of Coronary Bifurcation Lesions, Part I: Implanting the First Stent in the Provisional Pathway. The 16th Expert Consensus Document of the European Bifurcation Club. EuroIntervention. 2022;18:e362–e376.

30. Lassen JF, Albiero R, Johnson TW, Burzotta F, Lefevre T, Iles TL, Pan M, Banning AP, Chatzizisis YS, Ferenc M, et al. Treatment of Coronary Bifurcation Lesions, part II: Implanting Two Stents. The 16th Expert Consensus Document of the European Bifurcation Club. EuroIntervention. 2022;18:457–470.

31. Murasato Y, Mori T, Okamura T, Nagoshi R, Fujimura T, Yamawaki M, Ono S, Serikawa T, Nakao F, Shite J, et al. Efficacy of the Proximal Optimization Technique on Crossover Stenting in Coronary Bifurcation Lesions in the 3D-OCT Bifurcation Registry. Int J Cardiovasc Imaging. 2019;35:981–990.

32. Okamura T, Nagoshi R, Fujimura T, Murasato Y, Yamawaki M, Ono S, Serikawa T, Hikichi Y, Norita H, Nakao F, et al. Impact of Guidewire Recrossing Point into Stent Jailed Side Branch for Optimal Kissing Balloon Dilatation: Core Lab 3D Optical Coherence Tomography Analysis. EuroIntervention. 2018;13:e1785–e1793.

33. Onuma Y, Katagiri Y, Burzotta F, Holm NR, Amabile N, Okamura T, Mintz GS, Darremont O, Lassen JF, Lefevre T, et al. Joint Consensus on the Use of OCT in Coronary Bifurcation Lesions by the European and Japanese Bifurcation Clubs. EuroIntervention. 2019;14:e1568–e1577.

34. Onuma Y, Kogame N, Sotomi Y, Miyazaki Y, Asano T, Takahashi K, Kawashima H, Ono M, Katagiri Y, Kyono H, et al. A Randomized Trial Evaluating Online 3-Dimensional Optical Frequency Domain Imaging-Guided Percutaneous Coronary Intervention in Bifurcation Lesions. Circ Cardiovasc Interv. 2020;13:e009183.

35. Lee CH, Nam CW, Cho YK, Yoon HJ, Kim KB, Gwon HC, Kim HS, Chun WJ, Han SH, Rha SW, et al. 5-Year Outcome of Simple Crossover Stenting in Coronary Bifurcation Lesions Compared With Side Branch Opening. JACC Asia. 2021;1:53–64.

36. Tarantini G, Fovino LN, Varbella F, Trabattoni D, Caramanno G, Trani C, De Cesare N, Esposito G, Montorfano M, Musto C, et al. A Large, Prospective, Multicentre Study of Left Main PCI Using a Latest-Generation Zotarolimus-eluting Stent: the ROLEX study. EuroIntervention. 2023;18:e1108–e1119.

37. Foin N, Torii R, Alegria E, Sen S, Petraco R, Nijjer S, Ghione M, Davies JE, Di Mario C. Location of Side Branch Access Critically Affects Results in Bifurcation Stenting: Insights from Bench Modeling and Computational Flow Simulation. Int J Cardiol. 2013;168:3623–3628.

38. Takahashi H, Otake H, Shinke T, Murasato Y, Kinoshita Y, Yamawaki M, Takeda Y, Fujii K, Yamada SI, Shimada Y, et al. Impact of Final Kissing Balloon Inflation on Vessel Healing Following Drug-eluting Stent Implantation: Insight from the Optical Coherence Tomography Sub-study of the J-REVERSE Trial. J Cardiol. 2016;68:504–511.

39. Burzotta F, Lassen JF, Lefevre T, Banning AP, Chatzizisis YS, Johnson TW, Ferenc M, Rathore S, Albiero R, Pan M, et al. Percutaneous Coronary Intervention for Bifurcation Coronary Lesions: The 15(th) Consensus Document from the European Bifurcation Club. EuroIntervention. 2021;16:1307–1317.

40. Murasato Y, Kinoshita Y, Yamawaki M, Shinke T, Otake H, Takeda Y, Fujii K, Yamada S, Shimada Y, Yamashita T, et al. Efficacy of Kissing Balloon Inflation after Provisional Stenting in Bifurcation Lesions Guided by Intravascular Ultrasound: Short and Midterm Results of the J-REVERSE Registry. EuroIntervention. 2016;11:e1237–e1248.

41. Thiele H, Zeymer U, Thelemann N, Neumann FJ, Hausleiter J, Abdel-Wahab M, Meyer-Saraei R, Fuernau G, Eitel I, Hambrecht R, et al. Intraaortic Balloon Pump in Cardiogenic Shock Complicating Acute Myocardial Infarction: Long-Term 6-Year Outcome of the Randomized IABP-SHOCK II Trial. Circulation. 2019;139:395–403.

42. Thiele H, Zeymer U, Akin I, Behnes M, Rassaf T, Mahabadi AA, Lehmann R, Eitel I, Graf T, Seidler T, et al. Extracorporeal Life Support in Infarct-Related Cardiogenic Shock. N Engl J Med 2023;389:1286–1297.

43. Basir MB, Kapur NK, Patel K, Salam MA, Schreiber T, Kaki A, Hanson I, Almany S, Timmis S, Dixon S, et al. Improved Outcomes Associated with the use of Shock Protocols: Updates from the National Cardiogenic Shock Initiative. Catheter Cardiovasc Interv. 2019;93:1173–1183.

44. Numasawa Y, Sawano M, Fukuoka R, Ejiri K, Kuno T, Shoji S, Kohsaka S. Antithrombotic Strategy for Patients with Acute Coronary Syndrome: A Perspective from East Asia. J Clin Med. 2020;9:1963.

